# Deep-Learning Approaches to Identify Critically Ill Patients at Emergency Department Triage Using Limited Information

**DOI:** 10.1101/2020.05.02.20089052

**Authors:** Joshua W. Joseph, Evan L. Leventhal, Anne V. Grossestreuer, Matthew L. Wong, Loren J. Joseph, Larry A. Nathanson, Michael W. Donnino, Noémie Elhadad, Leon D. Sanchez

**Author notes:** Corresponding Author Address: Joshua W. Joseph, MD, MS, MBE, One Deaconess Road, Department of Emergency Medicine, Beth Israel Deaconess Medical Center Boston, MA 02215 USA. **Author Contributions:** JWJ, ELL, LDS, and NE conceived of the overall study design. Initial data gathering was performed by ELL and LAN, with model development and initial statistical analysis performed by JWJ. NE provided critical review of model design. AVG provided additional statistical review. JWJ drafted the initial manuscript, LJJ provided initial manuscript review. All authors contributed significantly to its revision. LDS and MWD provided supervision of the study. JWJ takes responsibility for the integrity of the data and the accuracy of the data analysis. **Funding/Support:** This work was conducted with support from CRICO - Risk Management Foundation of the Harvard Medical Institutions Incorporated (Improving Patient Safety Grant). **Role of the Funder:** CRICO - Risk Management Foundation of the Harvard Medical Institutions Incorporated, was not involved in the design and conduct of the study; collection, management, analysis, or interpretation of the data; or preparation and review of the manuscript. CRICO did not approve the manuscript and had no input in the decision to submit the manuscript for publication. No publication restrictions apply. The content is solely the responsibility of the authors and does not necessarily represent the official views of Harvard University and its affiliated academic healthcare centers.

## Abstract

**Importance:** Triage quickly identifies critically ill patients, helping to facilitate timely interventions. Many emergency departments use the emergency severity index (ESI) or abnormal vital sign thresholds to identify critically ill patients. However, both rely on fixed thresholds, and false activations detract from efficient care. Prior research suggests that machine-learning approaches may improve triage accuracy, but have relied on information that is often unavailable during the triage process.

**Objective:** We examined whether deep-learning approaches could identify critically ill patients using data immediately available at triage with greater discriminative power than ESI or abnormal vital sign thresholds.

**Design:** Retrospective, cross-sectional study.

**Setting:** An urban tertiary care hospital in the Northeastern United States.

**Participants:** Adult patients presenting to the emergency department from 1/1/2012 - 1/1/2020 were included. Deidentified triage information included structured data (age, sex, initial vital signs, ESI score, and clinical trigger activation due to abnormal vital signs), and textual data (chief complaint) with critical illness (defined as mortality or ICU admission within 24 hours) as the outcome.

**Interventions:** Three progressively complex deep-learning models were trained (logistic regression on structured data, neural network on structured data, and neural network on structured and textual data), and applied to triage information from all patients.

**Main Outcomes and Measures:** The primary outcome was the accuracy of the model for predicting whether patients were critically ill using area under the receiver-operator curve (AUC), as compared to ESI, utilizing a 10-fold cross-validation.

**Results:** 445,925 patients were included, with 60,901 (13.7%) critically ill. Vital sign thresholds identified critically ill patients with AUC 0.521 (95% CI 0.519 -- 0.522), and ESI less than 3 demonstrated AUC 0.672 (95% CI 0.671 -- 0.674), logistic regression classified patients with AUC 0.803 (95% CI 0.802 -- 0.804), neural network with structured data with 0.811 (95% CI 0.807 - 0.815), and the neural network model with textual data with AUC 0.851 (95% CI 0.849 -- 0.852).

**Conclusions and Relevance:** Deep-learning techniques represent a promising method of enhancing the triage process, even when working from limited information. Further research is needed to determine if improved predictions can be translated into meaningful clinical and operational benefits.

## Introduction

### Background

Triage quickly identifies critically ill patients, helping to facilitate rapid interventions with the goal of altering the course of disease. Many emergency departments (EDs) use the emergency severity index (ESI) or other standardized scores to facilitate triage and prioritize patients.(1) Concurrently, many EDs combine this with a clinical trigger system, which mobilizes available physicians and nurses to see patients with acute ESI scores or abnormal vital signs immediately after initial triage. The use of clinical triggers to mobilize clinicians has been demonstrated to improve patients’ time to physician evaluation and time to antibiotics.(2)

### Importance

Both ESI and vital sign triggers rely on specific vital sign thresholds. While ESI is among the most validated algorithms for triage, numerous studies have shown that both under-triage and over-triage remain persistent issues.(3,4) When patients are under-triaged, opportunities to change the course of disease are missed, while over-triage has the potential to disrupt physicians’ and nurses’ workflows, detracting from safe and efficient care for other patients in the ED. In particular, better understanding of the effects of advanced age, the influence of specific chief complaints, and more robust criteria for vital sign abnormalities have been highlighted as areas for improvement to the current ESI.(4,5) Studies examining machine-learning approaches have shown promise in supplementing the ESI score at triage, including random forest models to help differentiate outcomes for patients within ESI categories (6), gradient boosting algorithms to predict admission (7), and outcomes in specific conditions, such as mortality in sepsis (8).

However, many of these studies have leveraged information, such as past medical histories, that often is not immediately available at triage. In particular, patients who are making their first contact with an ED rarely bring medical history information in a ready electronic format. Patients also may be unable to meaningfully provide a clear past medical history, whether due to dementia, language barriers, or limited health literacy. Depending on their complexity, some machine-learning approaches are not readily integrated with commercial electronic health record (EHR) systems, and may require considerable effort to tune and adapt to a health system’s specific population.

Deep neural networks are a family of machine-learning algorithms which have led to rapid improvements across a variety of domains, including computer vision and natural language processing, and have made progress toward automated diagnosis in subfields of radiology and pathology. Compared to traditional methods of regression analysis, neural networks are intended to model multiple levels of complex, high dimensional interaction terms between independent variables without loss of specificity, and can be rapidly retrained to account for subtle differences between populations. In the last few years, several open source frameworks have made it simple to develop, deploy, and share these algorithms without the need for specialized equipment.

### Goals of This Investigation

We examined whether a set of progressively complex deep-learning algorithms could identify critically ill patients with greater discriminative power than ESI or vital sign triggers alone using information immediately available at triage, as measured by the area under the receiver-operator curve (AUC).

## Methods

### Study design and selection of participants

This was an observational study examining a retrospective cohort of adult patients who visited an academic, urban ED at a tertiary care center in the Northeastern United States with an average volume of 55,000 visits annually. All patients between 1/1/2012 and 1/1/2020 were screened for the study. Patients were included if their data included triage vital signs (patients with one or more vital signs were included, patients with none were not). ESI score, whether a vital sign trigger had been activated, and ultimate disposition (including whether they expired within the ED) were recorded for all patients. Clinical data was obtained from an automated quality assurance database for the ED information system. Data were de-identified during extraction using the HIPAA SAFE HARBOR method, and the study authors were blinded to identifying information. The study was structured with respect to the Transparent reporting of a multivariable prediction model for individual prognosis or diagnosis (TRIPOD) statement.(9) The host institutional review board granted an exemption for de-identified data used.

### Measurements

Vital signs were defined as a patient’s initial triage measurement of temperature, heart rate, systolic and diastolic blood pressure, respiratory rate, and percent oxygenation. To exclude potentially erroneous data entries, vital signs were included in the analysis provided they fell within broad physiologically feasible ranges, including temperature below 110 degrees, heart rate below 300 beats/minute, systolic and diastolic measurements below 300mmHg, respiratory rate below 80 respirations/minute, and oxygenation at or below 100%. Missing or spurious data were considered as null values for the analysis.

Triage chief complaints were included as free text entered immediately at a patient’s arrival at triage. Chief complaints were not standardized during the study period, and typographic errors and blank entries were included in the analysis for fidelity. Nursing documentation after the patient’s initial registration was not included. As a result of de-identification, patients older than 89 years of age were included within a single 90+ age category.

Vital sign trigger criteria were defined as heart rate less than 40 or greater than 130 beats/min, respiratory rate below 8 or above 30 respirations/min, systolic blood pressure below 90mmHg, or an oxygen saturation below 90% on room air, which is standard practice for trigger activations at triage for the institution.(2) As a result of the clinical system at our institution, all patients who meet clinical trigger criteria are classified as having an ESI score of 1 or 2.

### Outcomes

The primary outcome was whether a patient was critically ill, defined as whether they expired within 24 hours of arrival, required ICU admission from the ED, or were transferred from an inpatient ward to the ICU (or for an emergent procedure) within 24 hours of admission. The data abstraction process included the possibility of a patient being discharged and returning to the ED as critically ill within 24 hours. There have been many different measures of resource use and illness severity used across studies evaluating the efficacy of triage and machine-learning predictions, including specific diagnoses such as sepsis (8), admission (7), and overall mortality.(10) The composite of mortality and ICU admission is an appealing compromise metric, as it identifies a population that is more likely to require rapid intervention than the larger population of patients needing admission.

### Neural Network Model Creation and Derivation

Neural networks consist of a series of nodes known as neurons, which take input variables, apply an affine transformation to the inputs based on a set of weights, and yield an output based on whether a discrete threshold, known as the activation function, has been met. A loss function measuring the neuron’s output relative to the correct diagnostic labels is used to adjust the neuron’s weights repeatedly until loss has been minimized. Accordingly, logistic regression can be thought of as a single neuron with a sigmoid activation function.

Neural networks leverage multiple layers of interconnected neurons to model high-degree interaction effects. For instance, a neuron within a deep neural network layer might recognize specific combinations of elevated heart rate and temperature, while a separate neuron within the same layer might recognize separate thresholds for the same variables when associated with a different age range or set of chief complaints.

Our models were created in TensorFlow, an open-source framework for deep-learning.(11) The triage data was split between the structured vital sign data, and the chief complaint text data. The vital sign data was normalized and used as the input for a logistic regression, and for a two-layer deep neural network. A third, deep neural network combining both structured and text data likewise used the vital sign data as the input to a smaller two-layer deep neural network. The chief complaint text data was first embedded into a text vector and input into a Long Short-Term Memory network, a standard architecture for text processing and in clinical natural language processing.(12,13)

The outputs of these two subnetworks were then concatenated and used as the cumulative input to a densely connected layer, with a final logistic prediction layer. Hyperparameters of the model, such as the number of neurons per layer and the total number of layers in the overall model, were adjusted using random search and subsequently manually adjusted, with adaptive moment estimation for optimization.(14) Data was divided into an 80:10:10 training:validation:testing split to avoid overfitting, with 10-fold cross-validation.

### Analysis

We report the diagnostic accuracy of the methods evaluated in this study in terms of their sensitivity, specificity, accuracy, and area under the receiver-operator curve (AUC) with 95% confidence intervals, relative to the reference standard of critical illness abstracted from the medical record. Statistical analysis was carried out in Python 3.8 utilizing the SciPy and SciKit-Learn scientific and machine-learning libraries.(15,16)

Prior studies of machine-learning at triage have suggested that critically ill patients represent a fraction of the total patients seen within the ED, estimated as 2% of patients by Raita et al.(17) and Levin et al.(6), who examined a large random sample of patients from the National Hospital Ambulatory Medical Care Survey (NHAMCS) and the yearly volume of an urban ED, respectively. As a result of the high proportion of lower-acuity patients in the underlying population, we used the AUC as the primary measure of test accuracy for comparison between the existing triage models and the deep-learning models.

## Results

### Characteristics of the study subjects

From 1/1/2012 and 1/1/2020, 445,925 adult patients met inclusion criteria, detailed in Figure 1a. Patient demographic and vital sign characteristics are detailed in Table 1. 60,901 (13.7%) of patients met criteria for critical illness. Vital sign information contained a missing or spurious datapoint for 34,827 (7.6%) of patients, most commonly in the form of a missing temperature measurement at triage (24,872; 5.6%). Missing or spurious vital signs (500, <0.1%) or ages (104, <0.1%) were entered into the neural network as null values. The full enrollment process is described in Figure 1.

**Figure 1.**
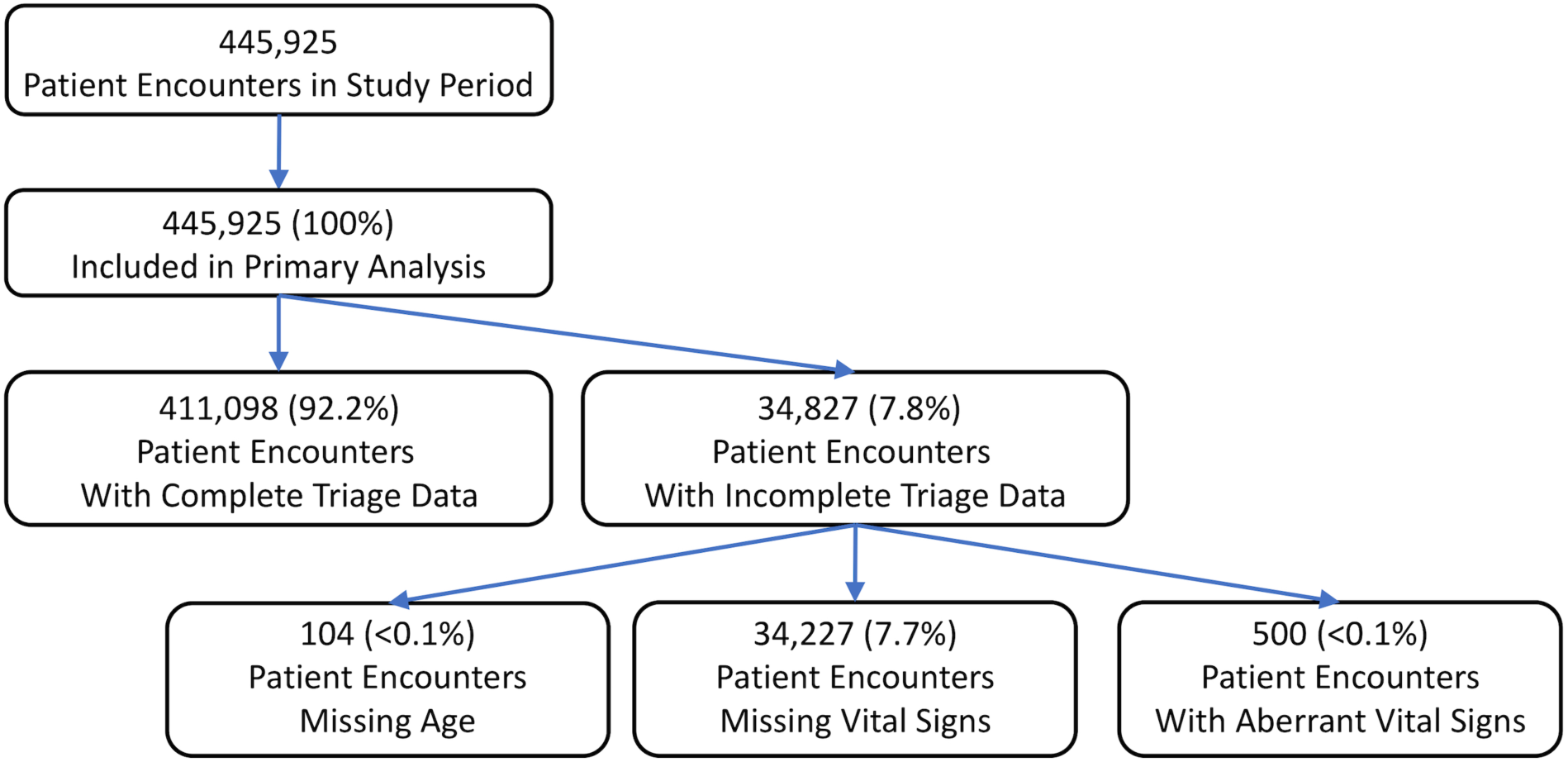
– Study Enrollment

**Table 1.**
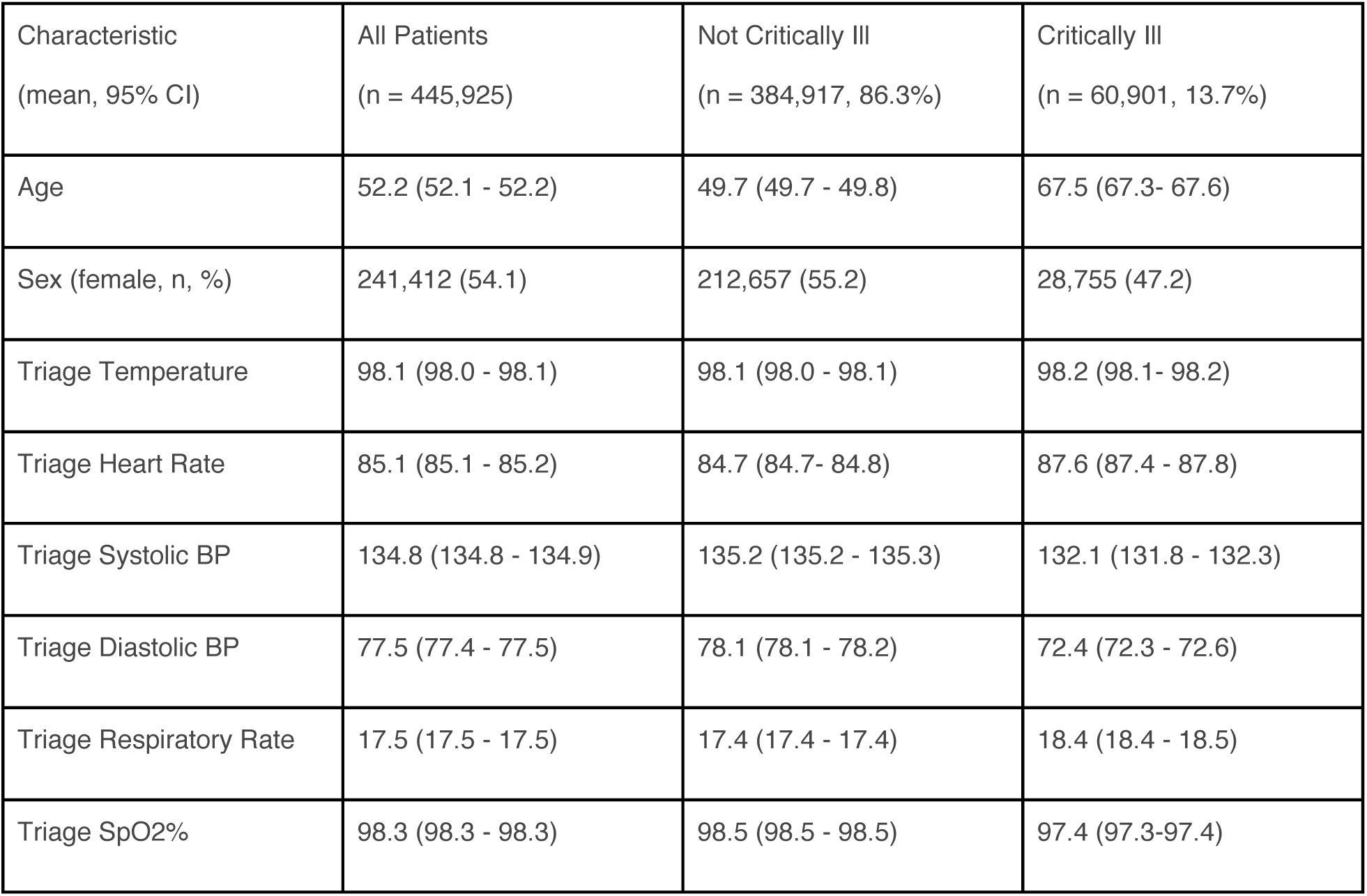
– Characteristics of the Study Participants

There were significant differences between the groups of patients assessed at triage who were critically ill and those who were not. Critically ill patients typically were older, more likely to be male, had faster heart rates and respiratory rates, lower blood pressures, and lower oxygen saturations. Admitted patients who were transferred to the ICU within 24 hours constituted a small portion of the overall population of the critically ill (n= 4,623; 7.6%). The full details of the population cohorts is detailed in Table 1.

### Main results

The existing standards of triage evaluation to identify critical patients at our institution, abnormal vital sign triggers and ESI scores less than or equal to two, demonstrated limited overall accuracy, and divergent sensitivity and specificity. Strict abnormal vital sign triggers demonstrated low discrimination (AUC 0.521, 95% 0.519 -- 0.522), very low sensitivity (0.050, 95% CI 0.050 -- 0.051) but very strong specificity (0.991, 95% CI 0.991 -- 0.991). Conversely, the ESI score demonstrated greater discrimination (AUC 0.697, 95% CI 0.696 -- 0.699), representing the product of significantly increased sensitivity but more modest specificity. The full details of the models’ diagnostic scores are presented in Table 2 and ROC curves in Figure 2.

**Table 2.** – Test Characteristics of the Diagnostic Approaches

| Method | Sensitivity (95% CI) | Specificity (95% CI) | AUC (95% CI) |
| --- | --- | --- | --- |
| Abnormal Vital Sign Trigger | 0.050 (0.050 -- 0.051) | 0.991 (0.991 -- 0.991) | 0.521 (0.519 -- 0.522) |
| Triage ESI <= 2 | 0.697 (0.696 -- 0.699) | 0.647 (0.646 -- 0.649) | 0.672 (0.671 -- 0.674) |
| Logistic Regression | 0.778 (0.775 -- 0.782) | 0.673 (0.669 -- 0.678) | 0.805 (0.801 -- 0.808) |
| Neural Network - Structured Data | 0.813 (0.811 -- 0.814) | 0.653 (0.652 -- 0.655) | 0.812 (0.811 -- 0.814) |
| Neural Network - Combined Data | 0.845 (0.844 -- 0.846) | 0.704 (0.702 -- 0.705) | 0.857 (0.856 -- 0.858) |

**Figure 2.**
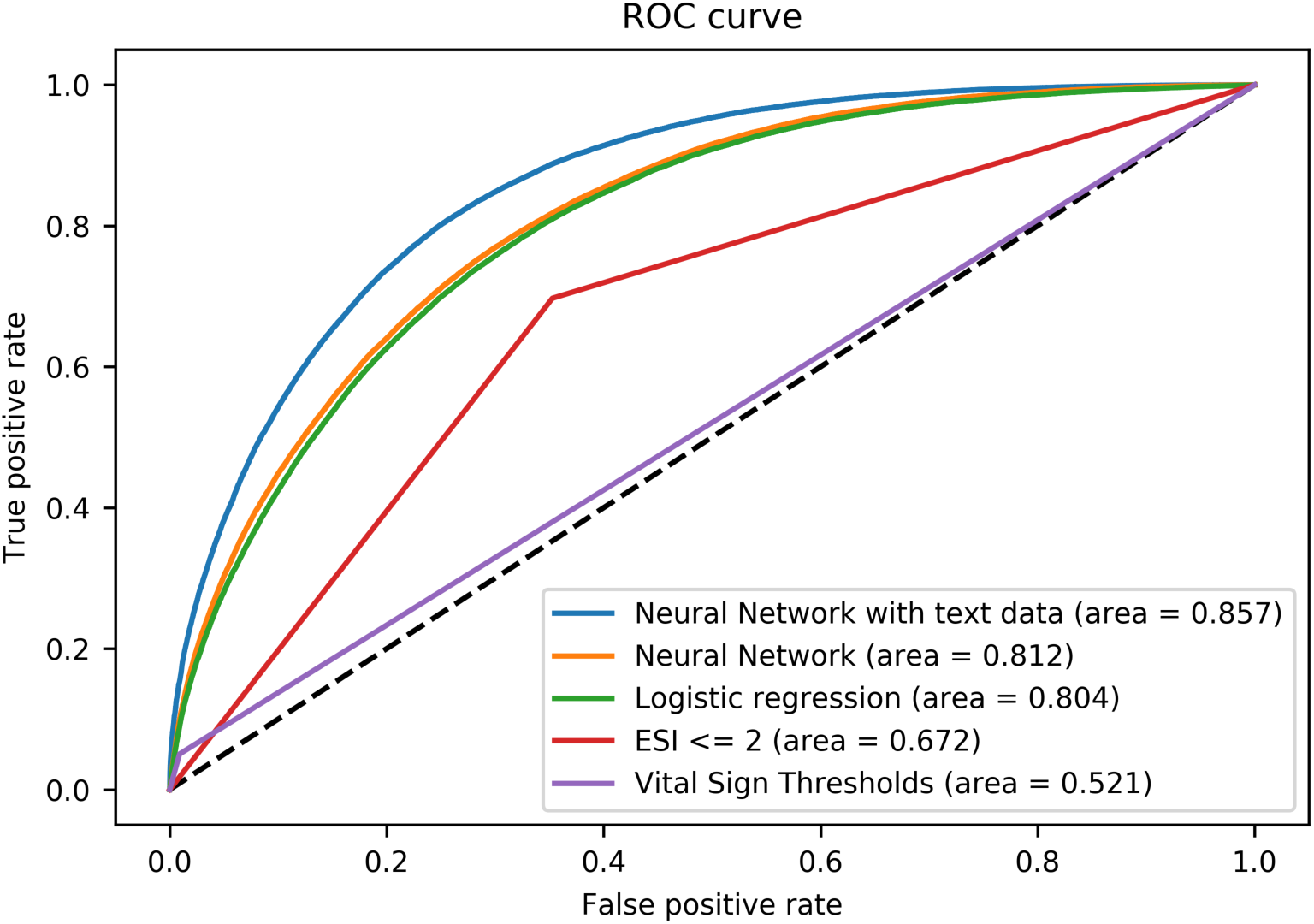
– Area under the Receiver Operator Curve for Detecting Critically III Patients by Analysis Method

The deep-learning approaches demonstrated progressive increases in sensitivity and AUC as the models became more complex. The design of the deep-learning models is illustrated in Figure 3a-c. The initial logistic regression on structured data yielded an AUC of 0.805 (95% CI 0.801 -- 0.808), with a sensitivity of 0.778 (95% CI 0.775 -- 0.782). The two-layer neural network on this same structured data demonstrated modest increases in AUC (0.812, 95% CI 0.811 -- 0.814) and sensitivity (0.813, 95% CI 0.811 -- 0.814).

**Figure 3a.**
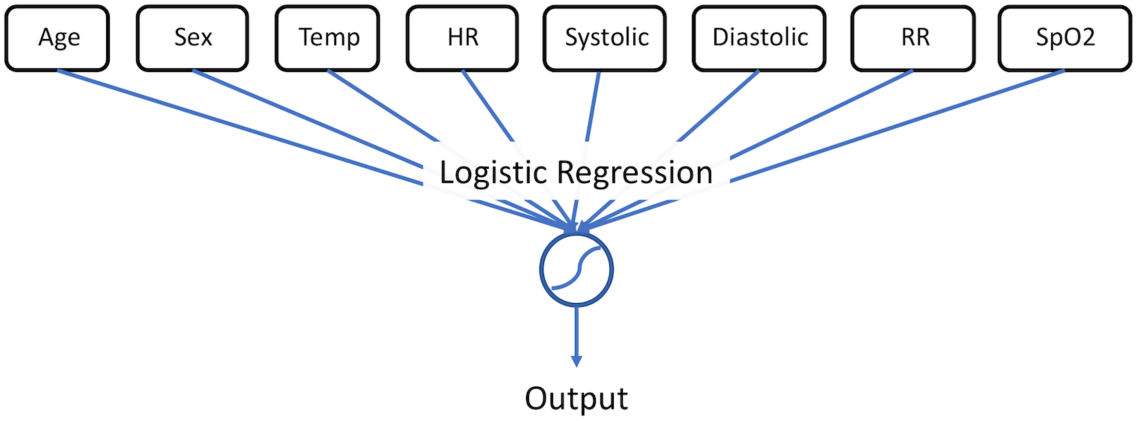
– Logistic Regression

**Figure 3b.**
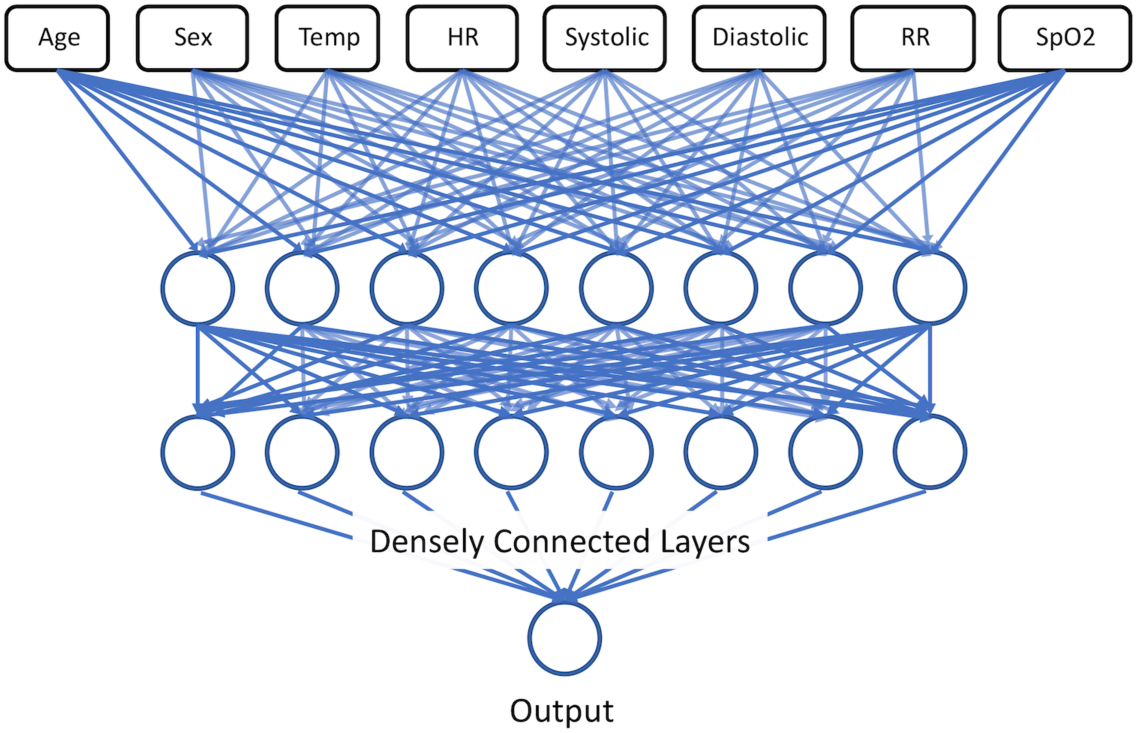
– Two Layer Dense Neural Network

**Figure 3c.**
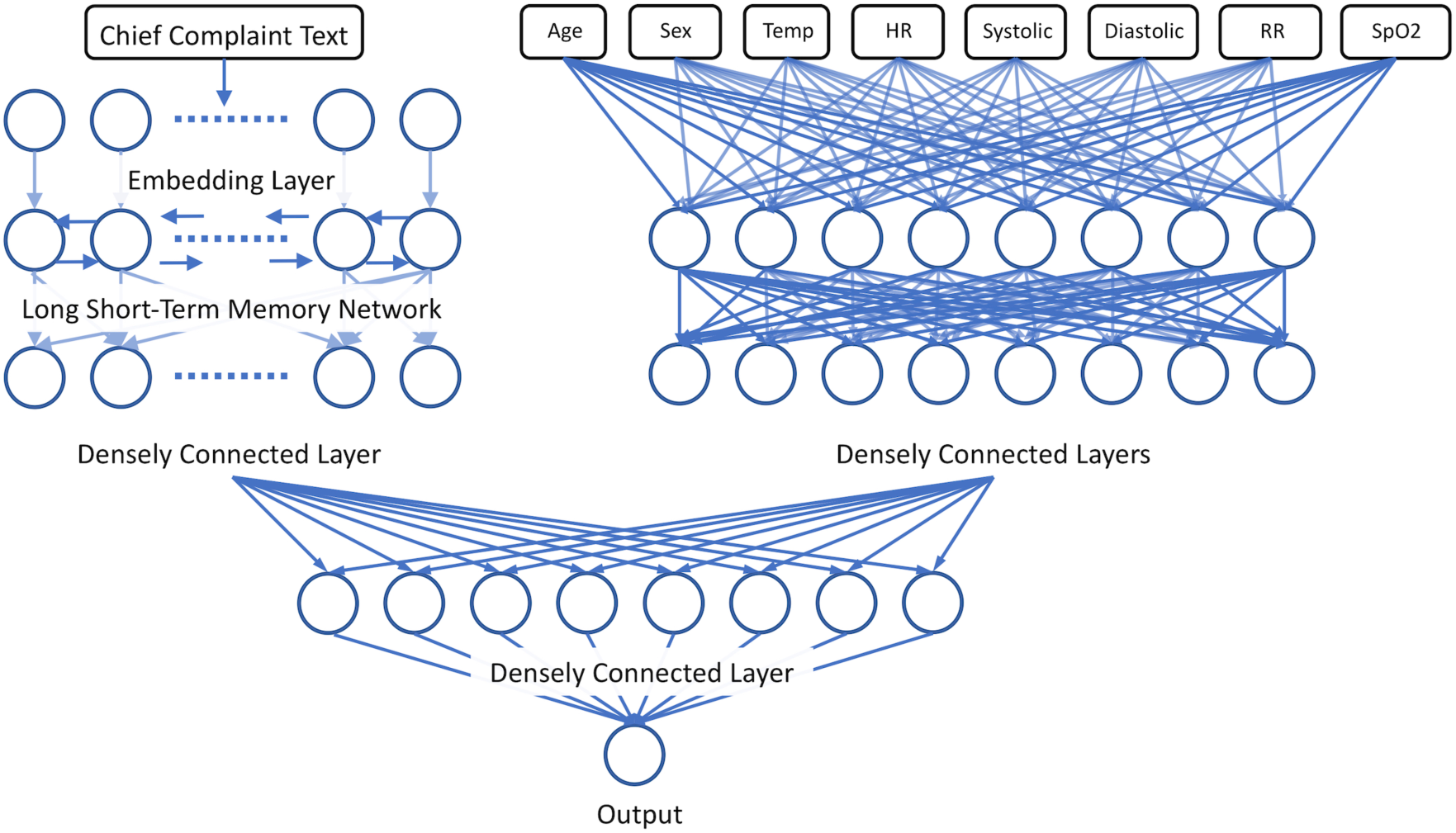
– Combined Neural Network

The addition of the unstructured chief complaint data provided further discriminatory power. After training and hyperparameter optimization, the final neural network model classified critically ill patients with AUC 0.851 (95% CI 0.849 -- 0.852), reflecting a total sensitivity of 0.845 (95% CI 0.844 -- 0.846).

## Discussion

Our study examines several models of triage which demonstrate increasing accuracy in tandem with increasing complexity. For those models relying on vital sign data alone, this progression is logical, as more complex models can draw more granular borders between data. The discrete vital sign cutoffs used for trigger criteria are simple to remember, and demonstrate considerable specificity, but miss a substantial number of critically ill patients. The enhanced accuracy of the more complex models likely reflects two factors - interaction effects between different vital signs, and the potential effects of age.

Early in their disease course, a critically ill patient may demonstrate subtle changes in multiple vital signs, which may be difficult to recognize individually, but meaningful in the aggregate. We are primed to recognize that a heart rate above 100 or a respiratory rate above 20 is abnormal because these numbers are salient.(18,19) Comparatively, a heart rate of 95 combined with a respiratory rate of 18 might be equally predictive of illness, which may be easy for a clinician to miss, but will not elude a regression. Similarly, there exist meaningful age-related variations within vital signs, particularly for the elderly, which the regression and neural network models can recognize, but might be lost on all but the most meticulous clinicians.(20,21) Cognitive aids exist for recognizing abnormal vital signs over broad age ranges, such as the Broselow tape in pediatric resuscitation (22), but no cognate tool exists for the adult and elderly populations. For the deep-learning models examined in our study, however, abnormal vital signs can be redefined to a patient’s age on a year-by-year basis.

The modest improvement in accuracy between the logistic regression model and the neural network model examining vital signs alone may reflect an underlying information-theoretic limit to a single set of measurements. Though both models are more accurate than rigid vital sign cutoffs and use of the ESI score, it is notable that optimal neural network model in our study was only two layers deep, and was not improved by adding additional layers representing higher-dimension interaction effects. This suggests that while vital signs are essential to the triage process, and their interpretation can be improved substantially, alone they are not sufficient to identify all critical patients.

Although the addition of textual chief complaint data entails only a small amount of additional data per patient, it was associated with significant improvements in both the sensitivity and specificity of the neural network model. This likely reflects the critical contextual information that a patient’s chief complaint provides. A young patient presenting with hypotension and tachypnea may not be critically ill if their complaint is anxiety, and the additional attention of being taken to a critical care bay might exacerbate their symptoms. But the same vital signs in a patient with a chief complaint of abdominal pain could be essential to identifying a ruptured ectopic pregnancy.

Our models demonstrate similar levels of accuracy to prior machine-learning approaches to predict admission from the ED. The neural network in our study achieved similar accuracy to that examined by Hong et al. which predicted the larger category of all patients requiring admission, and Levin et al., which additionally predicted a patient’s specific ESI score.(6,7) However, a significant distinction between our model and similar approaches, is that ours depends only on information immediately available at triage. The models examined by Hong et al. had the benefit of using the triage ESI score as an input variable. While predicting the larger population of patients requiring admission is important for operations management, and the triage ESI score represents a rich source of data, it represents a prediction that is informed by the triage process rather than informing it. Conversely, the e-triage system outlined by Levin et al. leverages pre-existing data within the medical record, which may disadvantage patients without prior access to care, or patients who cannot provide a history.(23) As a result of using an open source framework, the neural networks examined in this study can be adapted for use in an EHR or web-browser, and are available to interested researchers by request.

As with any decision support tool, it would be a mistake to use our model to substitute for the judgement of emergency physicians and nurses. Instead, our results suggest that a neural network model can be a powerful supplement to clinicians’ immediate evaluations, potentially alerting them to subtle but important changes in vital signs, which have special significance relative to patients’ ages and chief complaints, akin to an automated, finely-grained Broselow tape for adults.

## Limitations

This was a retrospective study conducted at a single urban tertiary care center, with a significantly higher proportion of critically-ill patients than has been reported in similar studies. This may be partly explained in terms of the relatively high average age of the patient population, but could also reflect institutional bias. Our composite outcome included transfers of admitted inpatients from the medical floor to the ICU within 24 hours; however, this represented a small portion of critically ill patients. Similarly, as abnormal vital signs may serve as an independent criteria for admitting patients to the ICU, the presence of abnormal vital signs may artificially enhance the accuracy of any predictive test of ICU admission, independently of underlying illness. Finally, due to the fact that our composite metric examines outcomes after care in the ED, a portion of patients who are triaged appropriately as critically ill, but who respond rapidly to treatment and ultimately do not need admission to an ICU, may be inappropriately mislabeled as not critically ill, artificially decreasing the accuracy of triage.

## Conclusions

In this single-center retrospective study of deep-learning approaches to identifying critically ill patients at ED triage, neural network models demonstrated significantly higher accuracy than traditional methods of triage, suggesting that these models can augment the triage process. While diagnosing patients who are critically ill more accurately can help to more appropriately mobilize resources within the ED to treat them, future studies are needed to assess the clinical and operational impact of using neural networks to enhance the triage process, and which critically ill patients can benefit most from rapid intervention.

## Data Availability

The source code of the analysis model is publicly available as a text supplement included with the manuscript, and for download as an online repository. Individual de-identified participant data is available on request in a modified format to accommodate HIPAA SAFE HARBOR criteria.

http://https://github.com/jwjoseph/NN_triage_predict

---

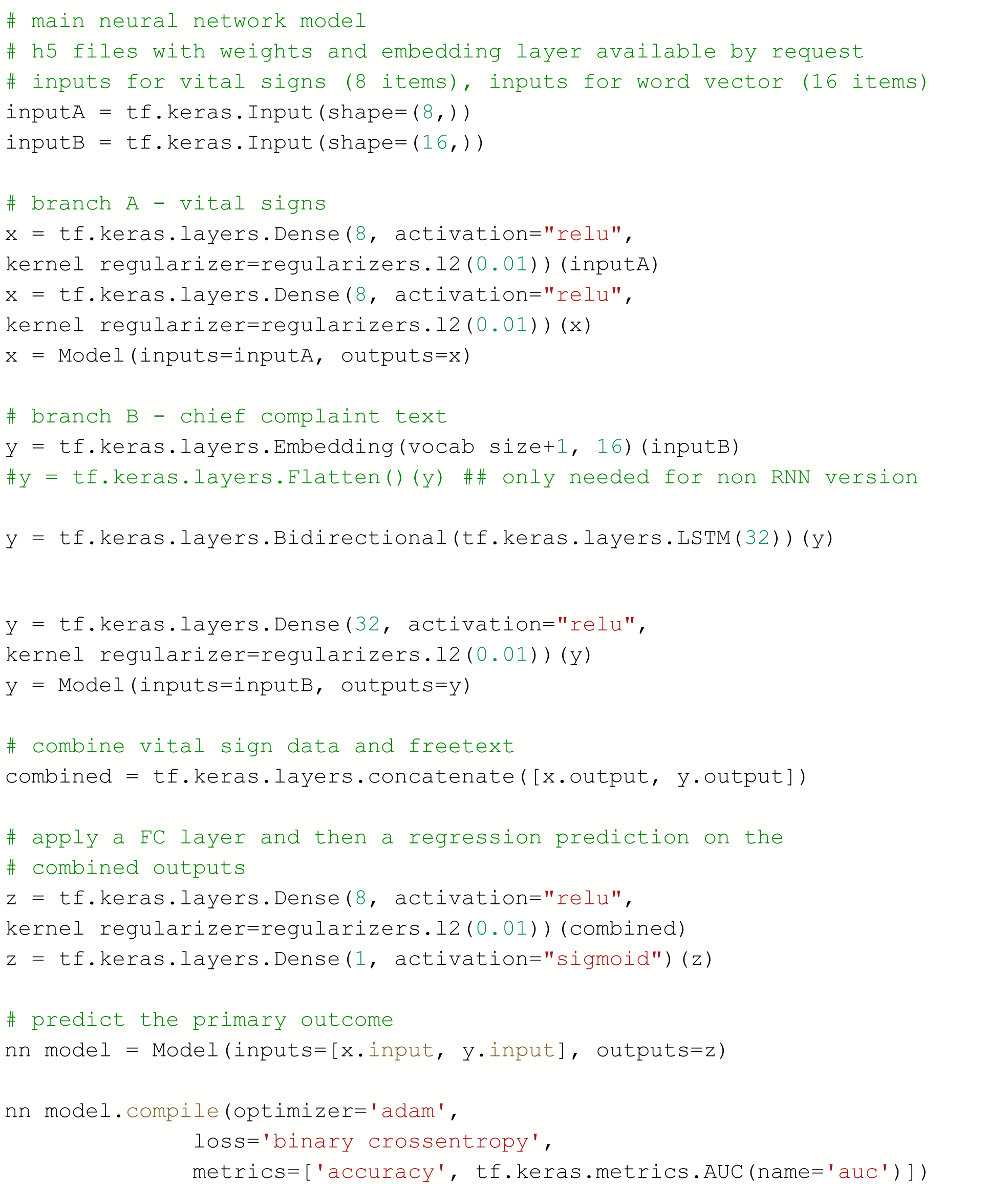

